# CPU-READY DEEP LEARNING APPROACH FOR ROBUST TISSUE REGION SEGMENTATION ACROSS MULTI-COHORT H&E AND IHC-STAINED WHOLE SLIDE IMAGES

**DOI:** 10.1101/2025.01.16.25320663

**Authors:** Sepideh Naghshineh Kani, Burak Can Soyak, Melih Gokce, Zeynep Duyar, Hasan Alicikus, Özlem Yapıcıer, Mustafa Umit Oner

## Abstract

With the rise of digital pathology, integrating digital slides with deep learning–based decision support systems is becoming increasingly common in clinical practice. Tissue region segmentation which is distinguishing tissue from background/artefacts, is an important pre-requisite in many digital pathology pipelines both for the laboratories as their first step in digitalizing the glass slides of tissue samples and turning them to whole slide images (WSIs) using scanners, and also for DL researches such as region-of-interest cropping, tumor detection, cell segmentation. However, it is well known that WSI scanners can fail in detecting all tissue regions, due to the tissue type, or due to weak staining and this is because of their not robust enough tissue detection algorithms which makes segmentation of WSIs a challenging task. Hence, this study develops a fast, lightweight, accurate, CPU-ready DL approach, enabling fast and reliable tissue region segmentation model by training and testing it across seven different institutional H&E and IHC stained WSIs to result a strong in generalization with the 22 to 56 s/WSI inference time using CPU that markedly outperforms classical OTSU thresholding, particularly in preserving challenging or faint tissue regions by achieving notably higher and more consistent performance than OTSU, with median Jaccard and Dice scores of approximately 0.86 and 0.92, respectively, compared to OTSU whcih was between 0.56 and 0.72. Our approach provides a practical, open-source solution for resource-limited pathology settings. We publicly released dataset obtained from Bahcesehir Medical School, and code to foster benchmarking and further advances in efficient, deployable computational pathology. The model could be used in digital slide scanners to improve the scanning process and in the pre-processing stages of DL pipelines to prepare high-quality datasets.

## 1 Introduction

Cancer is a leading cause of death worldwide, with one in five people developing it Bray et al. [2024]. Histopathology is an essential field in cancer studies, analyzing tissues at cell level. The samples obtained from patients, after undergoing several steps, are stained using various dyes and techniques to highlight specific structures or cell components within the tissue. Hematoxylin and Eosin (H&E) staining, for example, is commonly used to visualize cell nuclei (with Hematoxylin) and cytoplasm (with Eosin).

Traditional pathology in the clinic involves manual examination under a microscope, while digital pathology uses digital slide scanners to convert glass slides into high-resolution digital images called whole slide images (WSIs) Pantanowitz et al. [2018]. These images are stored in a pyramid structure where the base level contains the highest resolution image. Subsequent levels in the pyramid are downsampled versions of the base image, representing the slide at progressively lower resolutions. WSIs enable researchers to view and analyze the sample on a computer screen rather than using a traditional microscope. Additionally, WSIs can be processed by powerful deep learning models, which can support human pathologist and decrease their workload while providing more objective results Zarella et al. [2019].

Tissue region segmentation which in simple term is distinguishing tissue from background/artefacts is a foundational step in many digital pathology pipelines for DL research such as region-of-interest cropping(as a first step in any kind of DL research related to digital pathology), tumor detection, cell segmentation, and it is also the first step in digitalizing the glass slides of tissue samples and turning them to WSIs in laboratories and hospitals using scanners. Segmentation of histopathological whole-slide images is a challenging task that requires dedicated approaches. In medical image analysis, segmentation of input data often forms a crucial preprocessing step, e.g. to reduce the amount of input to the advanced and often computationally costly main processing. Classical techniques such as OTSU thresholding Otsu [1979], region growing Adams and Bischof [1994],morphological operations like watershed segmentation Vincent and Soille [1991], and edge-based methods Canny [1983] have been widely applied for this task each with strengths and limitations. While computationally efficient, they are highly sensitive to stain variability, background artefacts, and scanning inconsistencies. As shown in Fig. 1, the OTSU method, which is commonly embedded in commercial scanners, often fails to capture complete tissue regions or over-segments entire slides as tissue, producing inaccurate masks and unnecessary storage overhead, while in digital pathology, accurate tissue region segmentation in WSIs is essential for diagnostics, in a way that a missing tissue regions can lead to misdiagnosis, and false detection can increase processing times. One way to overcome this issue in laboratories is to annotate the tissue region manually which is time consuming because of the high work load and the other way is as mentioned before to scan the whole parts of the glass which will increase the storage and time consuming, but the same can not be applied in DL research for making tissue masks for the region of intereset (ROI) cropping. These shortcomings highlight the need for more reliable segmentation strategies in diverse staining conditions Vahadane et al. [2016], Tellez et al. [2019].

**Figure 1.**
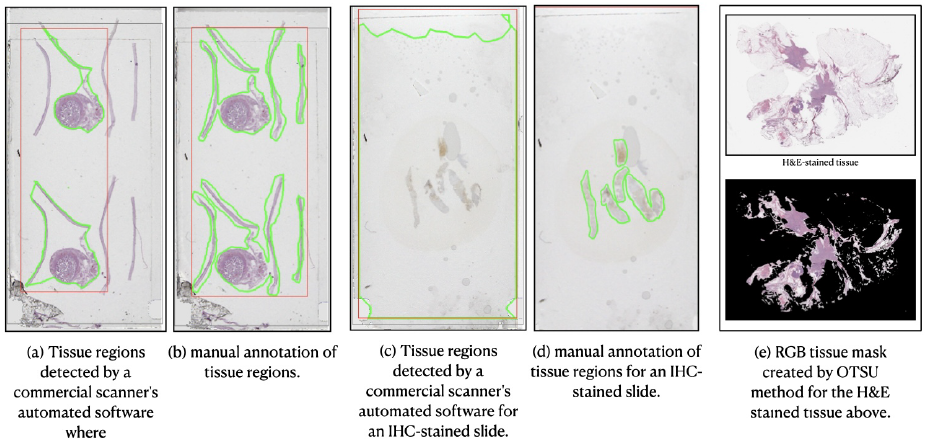
Problems with the OTSU method as a semi-automated classical segmentation method. (a) shows the tissue areas detected by a commercial scanner’s detection algorithm in an H&E-stained slide, where green contours highlight the tissue regions and red contours the region of interest (ROI), which are inaccurate. (b) shows the manually annotated tissue regions which are accurate. (c) shows that the scanner’s detection algorithm detected all pixels and regions as tissue areas causing unnecessary processing and using more storage in an IHC-stained slide. (d) shows the manually annotated tissue regions. (e) Original RGB image and masked tissue regions for machine learning dataset preparation.

Artificial intelligence (AI) and more specifically Deep Learning (DL) has shown remarkable performance in diagnostic and prognostic tasks in digital histopathology during the past decade. A recent systematic review and meta-analysis included diagnostic accuracy studies using any type of AI applied to over 150,000 whole slide images (WSIs) for any disease, explained that AI is having high diagnostic accuracy in the reported areas but requires more rigorous evaluation of its performance while these studies reported a mean sensitivity of 96.3% (CI 94.1-97.7) and mean specificity of 93.3% (CI 90.5-95.4) McGenity et al. [2024]. Other surveys similarly describe the contributions of deep learning to canonical tasks in medical image analysis: classification, detection, segmentation, registration, retrieval, image generation and enhancement. It is evident that deep learning has pervaded every aspect of medical image analysis Litjens et al. [2017], Srinidhi et al. [2021]. Another recent review highlights both the promise and practical pitfalls of deploying digital and AI-based pathology (DAIP); It is been concluded that AI demonstrates the potential to make a pathologist’s and institution’s workflow more efficient and accessible, but there are many other considerations to take into account, including ethical and legal issues as well as cost restrictions. Shen and colleagues [2025].

Despite the recent advantages of large Deep learning(DL) models in generalizing across diverse datasets, wide clinical adoption faces multiple obstacles: (1) variability in tissue sample taking, staining, and slide preparation adversely affect model robustness Pye et al. 2025, Vahadane et al. [2016], Tellez et al. [2019]; (2) Most of the DL models are too large for deployment and need GPU which is a drawback in resource-constrained settings Chen et al. [2016] and their high computational complexity also increases the risk of overfitting, especially with smaller datasets Zhang et al. [2021]; (3)

Most models are developed and validated only on data obtained from one center, which means it can not generalize across institutions; and (4) Although tissue region segmentation is a critical pre-processing step, but is often overlooked or handled by large and high computationally complex models with limited inference speed Campanella et al. [2019], Lu et al. [2021].

While many segmentation models have been developed and deployed for WSIs, efficient tissue region segmentation remains underexplored. There is a pressing need for tissue region segmentation models that have a balance between accuracy, efficiency, and robustness— particularly models small enough for deployment in typical pathology settings where GPU resources may be limited or non-existent and for deep learning researchers in digital pathology field in their data pre-processing step for creating accurate tissue masks especially for IHC-stained images. For satisfying this aim, in this study, we propose a lightweight CNN-based (modified LeNet5) tissue region segmentation model, designed for CPU-based inference on WSIs, with emphasis on cross-institutional generalization and integration into digital pathology pipelines which outperforming traditional methods and enhancing downstream DL model performance. Our key contributions are:

1. **Massive Annotated Data Collection:** We obtain the performance gain at the cost of many annotated examples. It is evident that deep learning algorithms are data voracious and demand millions of training examples. Collecting data, in general, is time-consuming, needs experts and is also expensive. Moreover, in medical imaging, it is not only about collecting annotations as they come from highly trained experts, but due to growing concerns on privacy, it is difficult to get the unlabeled examples Shamshad et al. [2023], Peng et al. [2021].
2. **Cross-Center Robustness:** A report in 2021 outlining factors needing to be addressed to close the ‘translation gap’ for AI applications in digital pathology emphasised that ‘variability in tissue acquisition and histopathology slide preparation could impact the performance of downstream image analysis tools’ and that ‘this variability is best accounted for during model development and validation before widespread adoption’ Steiner et al. [2021]. Starting from this point, we trained and validate the model across multi-institutional with different scanners and multi-stain datasets for different types of cancer, assessing generalization and to close the ‘translation gap’.
3. **Efficient Architecture Design:** We introduced a lightweight CNN model(modified LeNet-5), designed for efficient, real-time tissue segmentation in resource-constrained healthcare centers that achieves a high segmentation performance with minimal parameters and FLOPs.
4. **Practical Deployment Performance:** We demonstrate inference time and memory usage in a CPU-only environment suitable for real pathology lab deployment where each slide based on their size can be scanned in 22 - 56 seconds with a high accuracy.
5. **Open Resource Release:** We publicly released our code, model weights, and a curated IHC and H&E stained WSIs of breast cancer from Bahcesehir University to foster reproducibility and future research.

The rest of the paper is structured as followed. Section **??** provides an overview of related work in digital pathology and segmentation tasks in this domain and tissue region segmentation. Section 3 details the datasets used from seven different cohorts, the proposed lightweight CNN architecture, the training and validation schemes, and the evaluation metrics and the baseline. Section 4 presents both quantitative and qualitative experimental results, including ablation studies and cross-institutional validations. Section 5 discusses the key findings, highlights the model’s robustness. Finally, Section 6 summarizes the contributions of this study and its limitations, and outlines potential clinical applications and suggests directions for future research

## 2 Related Works

Deep learning models for segmentation tasks, such as deep convolutional neural networks (CNNs) Alzubaidi et al. [2021] like ResNet Targ et al. [2016], VGG Koonce [2021], Fully Convolutional Neural Networks (FCNNs) Long et al. [2015] and specialized architectures like U-Net Ronneberger et al. [2015], as well as architectures like UNet++ Zhou et al. [2020] which are used specifically for digital pathology images and HoVer-Net Graham et al. [2019], have shown remarkable performance in various image analysis tasks, including various segmentation tasks.

These models, by using their high computationally complex architectures, capture detailed patterns in WSIs, leading to high accuracy in diverse stained slides. For instance, Hossain et al. applied DoubleU-Net Jha et al. [2020] and ResUNet++ Jha et al. [2019] to tasks like artefact segmentation, where DoubleU-Net is basically a combination of two U-Net architectures stacked on top of each other and can be used as a strong baseline for both medical image segmentation and ResUNet++ is an improved ResUNet architecture for colonoscopic image segmentation Hossain et al. [2023]., and Bándi *et al*. applied FCNN and U-net CNN (UCNN) Hegde et al. [2018] method to tasks like foreground extraction in H&E and IHC stained tissues, comparing them with the Foreground Extraction from Structure Information (FESI) method Bándi et al. [2017]. These models by combining and utilizing feature extraction and attention mechanisms are able to capture complex tissue patterns, and achieve state-of-the-art segmentation and classification results in biomedical and digital pathology domain; comprehensive reviews of transformer-based medical imaging further contextualise these trends Shamshad et al. [2023]. However, despite the advantages of large DL models in generalizing across diverse datasets Szegedy et al. [2015], they require significant computational resources and have long inference time, making them not suitable for institutions with limited infrastructure and real-time clinical applications Chen et al. [2016]. These models’ high computational complex architecture also increases the risk of overfitting, especially in this domain (e.g., medical image analysis tasks) where it is challenging to find enough data and therefore most of the times models are trained with smaller datasets Zhang et al. [2021].

There is a growing attention among researchers to improve efficiency of deep learning models while reducing their complexity in medical image segmentation tasks. For example, Dinh *et al*. proposed U-Lite, a U-Net variant with just 878k parameters (35× fewer than typical U-Net) using depthwise separable convolutions, showing competitive performance on medical tasks Nguyen et al. [2025]. More recently, Zhang *et al*. combined a lightweight CNN (RepViT) with the Segment Anything Model (SAM) via knowledge distillation Gou et al. [2021] to achieve efficient segmentation in medical imaging contexts Zhang et al. 2024. There are also hybrid CNN– Transformer lightweight approaches such as MobileUtr tailored for efficient medical image segmentation Tang et al. 2023. In a broader review, Yao *et al*. categorise various architectures and highlight the trade-offs between segmentation accuracy and computational cost in medical domains Yao et al. 2024.

While the reviewed studies collectively represent significant progress in deep learning–based medical image segmentation, it should be noted that only one of them (Bándi *et al*.) focuses on tissue region segmentation. The rest mainly address different segmentation objectives such as nuclei, glands, or artefacts in WSIs. However, accurate and efficient tissue region segmentation is a crucial prerequisite for downstream analyses and model deployment. Therefore, our work contributes to the field by emphasising this fundamental yet overlooked step, proposing a lightweight and robust method that can accelerate and enhance the performance of subsequent segmentation tasks.

## 3 Materials and Methods

### 3.1 Datasets and Ethics

As mentioned in Section 1, we obtain the performance gain at the cost of many annotated examples; therefore we assembled a multi-institutional dataset of hematoxylin&eosin (H&E) and immunohistochemistry (IHC) whole-slide images (WSIs) from different types of cancer spanning public challenges and a private clinical cohort. All WSIs used from public sources were cited according to their licenses; private data (BAU) were de-identified prior to analysis. This study was approved by Bahçeşehir University Clinical Research IRB (document no: 2022-10/03). The BAU breast cohort (H&E and IHC) was released publicly at Zenodo and is accessible at https://zenodo.org/records/14131968.

### 3.2 Data Sources

#### TCGA

The Cancer Genome Atlas (GDC portal), which is a robust data-driven platform that allows cancer researchers and bioinformaticians to search and download cancer data for analysis. TCGA data are in SVS file format and are suitable-quality breast cancer National Cancer Institute.

#### CAMELYON17

Contains whole-slide images (WSI) of hematoxylin and eosin (H&E) stained lymph node sections and is collected from 5 medical centers in the Netherlands. WSIs are provided as TIFF images and are organized by the patient. Patients consist of 5 lymph nodes, and every slide holds sections of just one lymph node CAMELYON17 Grand Challenge.

#### HER2 Scoring (Nottingham 2016)

Since IHC-stained slides are one of the most problematic in digital histopathology studies, especially in making tissue masks using image processing techniques, training our model with this dataset was essential. This contest provided a dataset of nearly 100 whole-slide images (up to ∼ 100,000 *×* 80,000 px).By including the Her2 scoring contest dataset, which provides for both H&E and IHC stained slides, in our research, we aim to train our system on a more extensive dataset, allowing it to effectively predict and segment a broader range of tissue regions HER2 Scoring Contest, Tissue Image Analytics (TIA) Group, University of Warwick.

#### HEROHE (ECDP 2020)

Breast H&E WSIs scanned on a 3DHistech Pannoramic 1000 (MIRAX, `.mrxs`+ multi-`.dat`) ECPD2020 Grand Challenge.

#### Bahçeşehir Medical School (BAU)

Private breast H&E and IHC WSIs which has received Institutional Review Board approval (document no: 2022-10/03) from Bahcesehir University Clinical Research Institutional Review Board. released at Zenodo: https://zenodo.org/records/14131968 Naghshineh Kani et al. [2024a].

#### PANDA

Prostate H&E biopsies from two centers; included to test cross-organ generalization. It was important that our model could perform on other cancer types. Since with more than 1 million new diagnoses reported every year, prostate cancer (PCa) is the second most common cancer among males worldwide, resulting in more than 350,000 deaths annually, we improved our dataset by adding the PANDA dataset PANDA Grand Challenge [2022].

#### TTS Singapore (prostate)

H&E WSIs (40 *×*, 0.25*µ*m/px) of 99 prostate cancer patients (prostatectomy and core needle biopsies) at Tan Tock Seng Hospital, Singapore using Aperio AT2 Slide Scanner (Leica Biosystems) Oner et al. [2022].

### 3.3 Dataset Preparation

All the stages of dataset preparation, including data segregation, tissue mask generation, and patch cropping, are illustrated and shown briefly in Fig. [2] and described in detail below.

**Figure 2.**
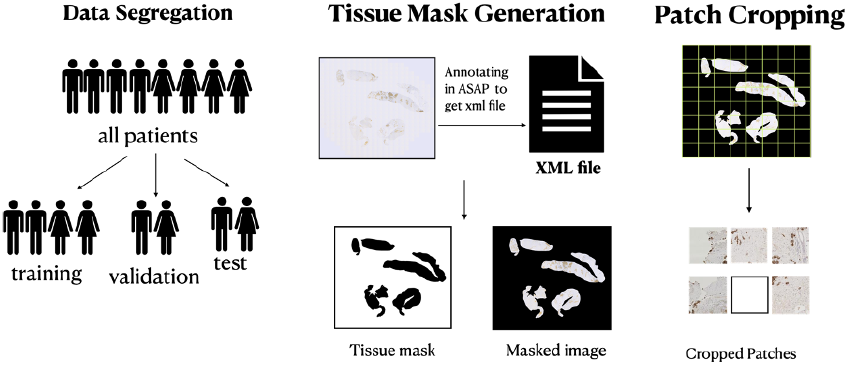
Dataset preparation

#### 3.3.1 Patient-level Splits

Data segregation is an important step in dataset preparation, where patients are divided into training, validation, and test sets. The training set trains machine learning models, while the validation set refines hyperparameters and chooses the best model. The model’s performance is then tested on unseen patients to simulate a real-world scenario. Data segregation at the patient level is crucial to avoid data leakage problems Oner et al. [2020], such as the correlation of tumor slides. Therefore, to avoid leakage (e.g., correlated slides from the same case), we split at the *patient level*. Each cohort was partitioned 70% training, 10% validation, and 20% testing. Slide overlap across splits was not permitted.

#### 3.3.2 Annotation and Reference Masks

Expert annotators delineated *tissue* vs. *background* using the ASAP tool, which is an annotation tool used primarily for image annotation to mark regions of interest within an image and associate them with specific labels or metadata, which stores human-readable XML polygon annotations and supports multi-vendor WSI formats. XML polygons were rasterized at the target pyramid level to produce binary reference masks. For quality control on tiny fragments, we applied a minimum area rule during labeling (Fig. [3]).

**Figure 3.**
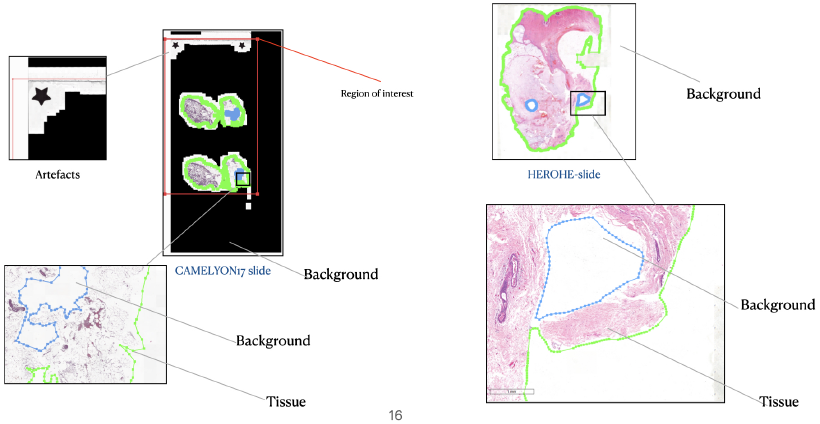
Tissue annotation is green, and background annotation is blue. Anything outside the tissue region is considered background. If the size of the tissue compared to the background is very small, to balance the classes, the tissue region and background are placed within a region of interest (ROI) box, which is red. Some WSIs have both black and white backgrounds.

#### 3.3.3 Tissue Mask Generating

The annotations made in ASAP Computational Pathology Group [2025] are typically stored in an XML file format, which includes information about the coordinates of the annotated regions on the image. These coordinates define the shape and location of the marked and annotated areas within the image. Using the obtained XML files, we created the tissue mask over a down-sampled version of the WSI, where the black areas of the binary tissue mask indicate tissue regions within the slide. In this way, we create labeled datasets for the model we are training, as shown in Fig. [4].

**Figure 4.**
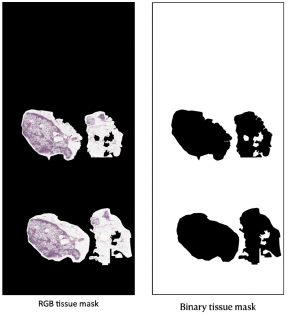
Created ground truth tissue masks on Level 5 with a Python code to parse the XML files for a WSI.

#### 3.3.4 Cropping in to Patches and Labeling

In this study cropping patches across tissue areas in WISs was the last stage in creating datasets. The reason for cropping the WSIs is their huge size, whereas most DL models work with small-sized images and cropping the WSIs significantly reduces the computational burden during training or analysis. Using a sliding window over the mask, patches are cropped from the desired resolution level of the slide using binary tissue masks as guides. Depending on the purpose, overlapping or non-overlapping sliding window methods may be used for cropping. During cropping, we label the tissue crops based on the hand-labeled mask. For each cropped region, if ≥10% of the patch area overlapped tissue and the mean RGB intensity was *<* 240*/*255 (to exclude near-blank glass), the patch was labeled, the patch was labeled *tissue*; otherwise *background*

#### 3.3.5 Resolution Patch Ablation

In this study we aimed to build a model that had minimum FLOPs and inference time; To achieve this, we cropped the WSIs at three different levels (4,5,6) with different patch sizes; such a cropped path at level 4 with patch sizes of 128*128 (dataset-L4-128) was referring to the same region in whole slide image, that a cropped patch at level 5 with patch sizes of 64*64 (dataset-L5-64) and a cropped patch at level 6 patch sizes of 32*32 (dataset-L6-32). In the same manner a patch from a specific region at level 4 with patch sizes of 64*64 (dataset-L4-64) was looking at the same region in whole slide image as a patch at level 5 with patch sizes of 32*32 (dataset-L5-32) and at level 6 patch sizes of 16*16 (dataset-L5-16). Eventually, the cropped patches will change into tensors and be normalised based on the mean and standard of the training set patches. After doing a set of data augmentation, we pushed the data into our DL model. By doing experiments on these different levels and patch sizes, we eventually founded the optimal path level and patch size for training the model which has the best minimum FLOPs and best test metric results trade off.

| Pyramid level | Patch size A | Patch size B |
| --- | --- | --- |
| Level 4 | $128 \times 128$ | $64 \times 64$ |
| Level 5 | $64 \times 64$ | $32 \times 32$ |
| Level 6 | $32 \times 32$ | $16 \times 16$ |

Based on validation performance and runtime, we selected **Level 4 with** 128 *×* 128 patches (L4_128) for all primary results.

### 3.4 Deep Learning Model

We adapted **LeNet-5** for 3-channel RGB input and the above patch sizes. LeNet-5’s compact design (convolutions, pooling, and fully connected heads) was chosen to minimize FLOPs and enable fast CPU-based inference and potential embedded deployment. FLOPS stands for Floating Point Operations Per Second. In the context of DL models, it refers to the computational complexity of the model, specifically the total number of arithmetic operations required for a single forward pass. Higher FLOPS implies more complex calculations and, therefore, higher resources (like time and energy) needed to run the model. To calculate the models FLOPs and their computational efficiency to estimate the model performance, we used a Facebook library named fvcore for this task. fvcore is the first tool that can provide both operator-level and module-level flop counts together and contains a flop-counting tool for PyTorch models fvc. We conducted a small resolution patch ablation and then fixed a single operating point for the main experiments. As shown in Table [2] by using `fvcore`, we estimated model computational complexity across input sizes; the final L4_128 configuration is ∼ 19.8 M FLOPs, whereas the smallest variant (e.g., 16 *×* 16 input) is ∼ 0.31 M FLOPs. (Exact counting scripts and configs are in the repository)

**Table 1.**
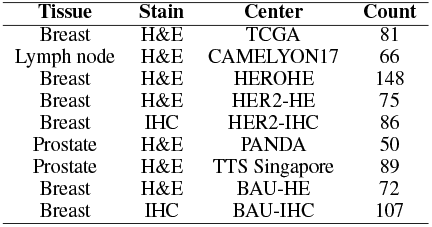
Overview of the development cohorts by tissue, stain, and source center. Values are the number of whole-slide images (WSIs). Data splits are at the *patient* level (70% train, 10% validation, 20% test) with no patient overlap across splits.

**Table 2.** Output of fvcore library for LeNet5 for RGB channel and input size of 128*×*128.

| [HTML]B7B7B7 Model | Total FLOPs | FLOP Table |  |
| --- | --- | --- | --- |
| LeNet5_128 | 19,179,528 | module | parameters or shape / flops |
|  |  | model | 1.979M / 19.18M |
|  |  | conv1 | 0.456K / 7.373M |
|  |  | conv1.weight | (6, 3, 5, 5) |
|  |  | conv1.bias | (6,) |
|  |  | conv2 | 2.416K / 9.83M |
|  |  | conv2.weight | (16, 6, 5, 5) |
|  |  | conv2.bias | (6,) |
|  |  | fc1 | 1.966M / 1.966M |
|  |  | fc1.weight | (120, 16384) |
|  |  | fc1.bias | (120,) |
|  |  | fc2 | 10.164K / 10.08K |
|  |  | fc2.weight | (84, 120) |
|  |  | fc2.bias | (84,) |
|  |  | fc3 | 0.17K / 0.168K |
|  |  | fc3.weight | (2, 84) |
|  |  | fc3.bias | (2,) |

### 3.5 Training Setup

The models were trained on two NVIDIA RTX A5000 GPUs, each with 24 GB of memory, on a Linux Ubuntu system. Before training, patches were converted to tensors, normalized by the training-set mean and standard deviation, and augmented to increase robustness to scanner and stain variability. Data augmentation included horizontal and vertical flips, rotations, and `ColorJitter`hue perturbations sampled uniformly in the range [™0.4, 0.4].

The model training was conducted in two phases. **Phase 1** involved experiments to identify the optimal patch size and magnification level that capture sufficient histological detail for accurate classification. In **Phase 2** We performed external validation for each model trained on data from individual institutions using datasets from other institutions that shared the same staining method and cancer type. For instance, H&E-stained whole-slide images (WSIs) of breast cancer from the BAU dataset were externally validated on H&E-stained WSIs of breast cancer from the TCGA and Her2-HE datasets, among others. **Phase 3** consisted of training the final model using the optimal patch size and magnification level by combining the data obtained from different institutions, that models trained on them showed better performance among other. then we compared the performance with OTSU thresholding.

We trained our model with batch size 1024, learning rate 10^™4^, **cross-entropy** loss, and **SGD** with momentum and L2 weight decay. We used Cross-Entropy loss or log loss as a loss/cost function to optimise the model during training in our classification model, whose output is a probability value between 0 and 1 and generally calculates the difference between two probability distributions. In binary classification, cross-entropy is calculated as:

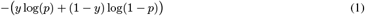

Where log is the natural logarithm, y is the binary indicator (0 or 1) for the class label, and p is the predicted probability. Since the objective is almost always to minimize the loss function and the lower the loss, the better the model is, using optimizers is essential, which are one of the most used methods for changing a model’s parameters to reduce a cost function in machine learning projects. For this matter, we used Stochastic gradient descent (SGD), a variant of the gradient descent algorithm used for optimizing machine learning models, and it addresses the computational inefficiency of traditional gradient descent methods when dealing with large datasets in machine learning projects. SGD randomly picks one data point from the whole data set at each iteration to reduce the computations enormously. We used momentum to accelerate the gradient vectors in the right direction in SGD to lead to a faster converging. In the context of training models, overfitting occurs when a model learns the underlying patterns in the data and captures noise and irrelevant details, making it perform poorly on new, unseen data. To prevent overfitting, we used weight decay (also known as L2 regularization) in the SGD optimizer, and it works by adding a penalty term to the loss function during training which aims not only to minimize the error between the predicted output and the actual target but also to minimize the magnitude of the weights.

To preserve the original LeNet-5 simplicity, we did not add batch normalization or dropout. We trained up to **1000 epochs**; the best checkpoint per experiment was selected using the validation metrics recorded each epoch. (Training/logging and analysis scripts— AUC, F1, precision/recall, confusion matrices, CI plots—are included in the repo.) We didn’t use early stop because during doing the experiments in Phase 1, we realized that training in high epochs is a must where the CNN is small.

in phase 2, we compared the performance of LeNet5 trained on different cohorts by doing an external validation on a combination of datasets we had and compared their performance using the F-1 score. In phase 3, by ensuring we include every kind of origin and stain type, we made a new dataset combining the patches from Camelyon17, HER2-IHC, HER2-HE, HEROHE, and TTS-Singapore and trained the model with this new dataset and did the external validation on the dataset from institutions we did not used for training this model and compared the results of this external validation with OTSU results for the same WSIs used for external validation.

### 3.6 Inference

In the segmentation task we will make binary tissue masks and for convenience in downstream pipelines, the tool can also export tissue-region polygons as XML alongside the binary mask (see Fig. [8]) which can be used in annotation tools like ASAP or Qupath QuPath Docs Authors [2025]. We performed sliding-window prediction over the WSI at L4_128 and mapped tile probabilities back to slide coordinates to obtain a binary tissue mask; minimal morphology (e.g., small hole filling) was applied when necessary. The released inference tool also exposes optional *hard* or *soft* voting across overlapping tiles; unless explicitly stated, main results do *not* use voting. On our setup, a full-slide segmentation typically completes in ∼22 **s/WSI**.

### 3.7 Classical Baseline (OTSU)

Over time, various image segmentation algorithms and techniques have been developed using domain-specific knowledge to address specific applications like medical imaging, automated driving, video surveillance, and machine vision. Each method has advantages and disadvantages. The threshold method, which works best with high-contrast pictures, separates an image into foreground and background areas depending on the threshold. Still, many details can get omitted during thresholding, and threshold errors are common. The process of generating a binary mask involves basic image processing operations over a down-sampled version of a WSI, including an OTSU thresholding, morphological operations, and median filtering to eliminate noise.

### 3.8 Evaluation

While accuracy is often the first metric to evaluate, it can be misleading in imbalanced datasets and in such scenarios, precision, recall, and F1 score provide deeper insights Hicks et al. [2022]. Also the Dice score and Jaccard index are commonly used metrics for the evaluation of segmentation tasks in medical imaging Bertels et al. [2019]. We report overlap metrics —Dice and Jaccard/IoU— computed against the binary reference masks, alongside precision and recall. For the quantitative contrast to OTSU, we evaluated on the external test set (70 images across tissue types with different staining methods and cancer types, different unseen institutions) and summarize results at slide and cohort level. The Jaccard Index and Dice Coefficient are similarity metrics used in various fields, particularly in data analysis, to compare the similarity and dissimilarity between sets or sequences.

### 3.9 Reproducibility

All code for data preparation (mask generation, cropping), training, evaluation, and inference— including configuration files and example commands— is released in a public repository and it has been deposited at Zenodo under https://zenodo.org/records/14294426 and is publicly available Naghshineh Kani et al. [2024b]. The repository provides a detailed step-by-step explanation, from preparing the ML dataset and training of tissue segmentation models to inference with the trained model, used to generate the masks in Figs. [8] and [7].

## 4 Results

### 4.1 Ablation: Resolution and Patch Size

In the first phase of experiments, we trained models on datasets obtained from each institution to find the optimal patch size and magnification level that capture sufficient details in WSIs for accurate classification. All datasets achieved high accuracy and AUC values above 0.98 and models trained on L4-128 was showing the highest among others. Since the AUC values were so close, it would be a shallow decision to identify the optimal operating-point choice as level 6 and patch size 16 *×* 16, for this reason we did a comparative evaluation using AUC confidence intervals via bootstrapping confirmed L4-128’s superiority, and the McNemar’s statistical test further validated significant differences (*p <* 0.05) between L4-128 and the other configurations (L5-64 and L6-32) 3. These findings established L4-128 as the optimal setting for subsequent experiments.

For instance, in TCGA (H&E) dataset, **L4_128** achieved F1 =**0.9770** and AUC =**0.9914**, exceeding L5_64 (F1 = 0.9724, AUC = 0.9908) and L6_32 (F1 = 0.9741, AUC = 0.9898). McNemar’s tests indicated significantly different error profiles: L4_128 vs. L5_64 (*p* = 9.59 *×* 10^™24^), L4_128 vs. L6_32 (*p* = 9.30 *×* 10^™9^), and L5_64 vs. L6_32 (*p* = 7 *×* 10^™4^). CI boxplots showed the narrowest spread for L4_128 (Fig. [5]).

**Figure 5.**
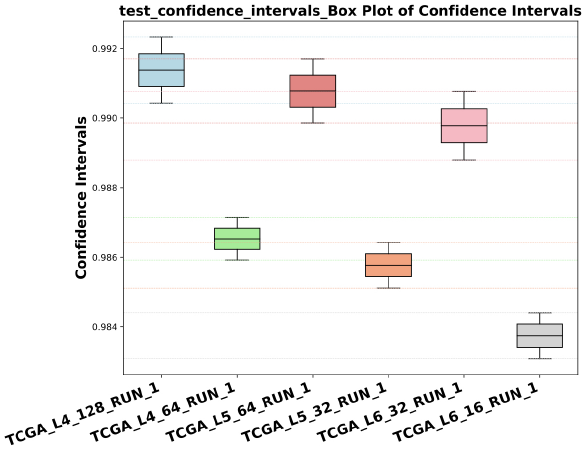
TCGA ablation: slide-level CI boxplots (F1) for L4_128, L5_64, L6_32; McNemar *p*-values: L4_128 vs. L5_64 = 9.59 *×* 10^™24^, L4_128 vs. L6_32 = 9.30 *×* 10^™9^, L5_64 vs. L6_32 = 7.0 *×* 10^™4^.

For other cohorts, where full ablation tables were not shown in this paper, McNemar tests and CI boxplots still pointed to L4_128 as a safe default: PANDA Karolinska: L4_128 vs. L5_64 (*p* = 0.6875), L4_128 vs. L6_32 (*p* = 0.125), L5_64 vs. L6_32 (*p* = 0.039). BAU_HE: all comparisons significant (e.g., L4_128 vs. L6_32 *p* = 3.86 *×* 10^™9^). HER2_HE: L4_128 better than L5_64 (*p* = 2.0 *×* 10^™4^) and L6_32 (*p <* 10^™4^); L5_64 vs. L6_32 not significant (*p* = 0.377). HER2_IHC: L4_128 ≈ L5_64 (*p* = 0.252), both ≫ L6_32 (*p <* 10^™4^). BAU_IHC: all pairs significant (e.g., L4_128 vs. L5_64 *p* = 2.08 *×* 10^™8^; L4_128 vs. L6_32 *p* = 1.96 *×* 10^™14^).

Given the consistent superiority of **L4_128** and corroborating McNemar/CI evidence on datasets we conducted all subsequent experiments on those cohorts *only at Level 4 with* 128 *×* 128 *patches*. The concise ablation table appears in Table [4].

**Table 3.** McNemar’s test results showing significance between models via p-values.

| Model1_vs_Model2 | Correct-Correct | Correct-Wrong | Wrong-Correct | Wrong-Wrong | p-value |
| --- | --- | --- | --- | --- | --- |
| TCGA_L4_128_vs_TCGA_L5_64 | 31,382 | 255 | 76 | 804 | 9.59E-24 |
| TCGA_L4_128_vs_TCGA_L6_32 | 31,403 | 234 | 125 | 755 | 9.3E-09 |
| TCGA_L5_64_vs_TCGA_L6_32 | 31,286 | 172 | 242 | 817 | 0.0006778 |

**Table 4.**
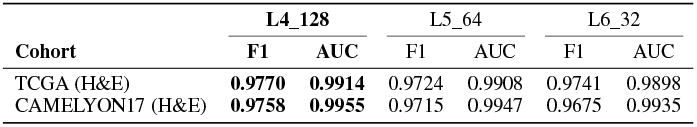
Ablation : F1 and AUC for TCGA and CAMELYON17 cohorts. Bold = best per row.

### 4.2 External Validation

It is crucial for DL models trained for histopathology tasks to be validate by completely unseen data from other cohorts to make sure the model will show the same performance on real-world data. For thus, we evaluated generalization on cohorts from other institutions we were having, using models trained at the selected operating point (L4_128).

Based on the experiments, we made a new dataset with the data of those models that showed better results in H&E-stained and IHC-stained breast cancer and H&E-stained prostate and lymph node biopsies on level 4 with a patch size of 128 and it included : HEROHE, Her2-IHC, Her2-HE, Camelyon17, and TTS-Singapore. This *final* model which is trained on the explained combined data from different cohorts,attained **AUC = 0.9988** with accuracy = 99.01%, F1 = 0.9542, and recall = 0.9804. The external validation set for the *final* model was a combination of TCGA, PANDA, and BAU-IHC and BAU-HE (Combined ext.).

As expected, performance metrics varied with class balance and foreground sparsity. All performance metrics (F1/precision/recall/accuracy) below are *uncalibrated* and computed at a fixed decision rule *p* ≥ 0.5 (no site-specific tuning).

Table [6] shows a characteristic pattern where ranking quality remains high (AUC near ceiling) but F1 drops under the fixed rule *p*≥ 0.5. For example, model trained on *BAU_IHC_L4_128* → and externally validated on HER2-IHC yields AUC = 0.9636 with F1 = 0.6766, and model trained on *PANDAS_Karolinska_L4_128* → which is externally validated on TTS-Singapore yields AUC = 0.9994 with F1 = 0.3126. This discrepancy is consistent with prevalence/threshold mismatch under domain shift. In practice, a small site-specific validation split can be used to select a threshold *t*^∗^ that maximizes F1, or to learn a simple post-hoc probability calibration (isotonic/beta) followed by a fixed threshold; both typically restore F1 with negligible overhead. We report uncalibrated results here and leave calibration as a deployment step.

**Table 5.** Internal performance at the selected operating point (L4_128).

| Cohort | F1 | Recall | Precision | AUC |
| --- | --- | --- | --- | --- |
| TCGA (H&E) | 0.9770 | 0.9659 | 0.9883 | 0.9914 |
| CAMELYON17 (H&E) | 0.9758 | 0.9598 | 0.9924 | 0.9955 |
| HEROHE (H&E) | 0.9733 | 0.9846 | 0.9622 | 0.9995 |
| TTS Singapore (H&E) | 0.9460 | 1.0000 | 0.8976 | 0.9994 |
| Combined (multi-cohort) | 0.9619 | 0.9689 | 0.9550 | 0.9984 |

**Table 6.** External validation summary (models trained at L4_128).

| Source model | External set | Accuracy (%) | AUC | F1 |
| --- | --- | --- | --- | --- |
| HER2_HE_L4_128 | TCGA / HEROHE / BAU-HE | 98.88 | 0.9398 | 0.9461 |
| TCGA_L4_128 | HER2-HE / HEROHE / BAU-HE | 98.57 | 0.9778 | 0.8928 |
| BAU_HE_L4_128 | TCGA / HEROHE / HER2-HE | 95.50 | 0.9568 | 0.8141 |
| HEROHE_L4_128 | TCGA / HER2-HE / BAU-HE | 91.57 | 0.9724 | 0.8956 |
| HER2_IHC_L4_128 | BAU-IHC | 97.84 | 0.9793 | 0.9400 |
| BAU_IHC_L4_128 | HER2-IHC | 92.50 | 0.9636 | 0.6766 |
| SG_L4_128 | PANDA Karolinska | 99.07 | **0.9996** | 0.9698 |
| PANDAS_Karolinska_L4_128 | TTS Singapore | 21.57 | 0.9994 | 0.3126 |
| **Final Model** | **Combined ext.** | **99.01** | **0.9988** | **0.9542** |

Note. F1/precision/recall computed at a fixed threshold $p \geq 0.5$ without site-specific calibration; AUC is threshold-independent.

In third phase of our experiments, for the **Final** model, the training accuracy was 98.38%. The validation accuracy was 99.54%, while the test accuracy reached 99.08%. The validation and test AUC scores were 0.9989 and 0.9984, respectively.

Figure [6] shows the comparison of segmentation performance between the conventional OTSU thresholding method and the proposed tissue region segmentation model. The boxplots of Jaccard and Dice coefficients show that our model achieved notably higher and more consistent performance than OTSU, with median Jaccard and Dice scores of approximately 0.86 and 0.92, respectively, compared to OTSU’s 0.56 and 0.72, indicating roughly 30% and 20% improvements and substantially reduced variability across samples; this performance gap would be even greater if the comparison were conducted solely on IHC-stained images, where OTSU typically struggles with faint tissue regions and background suppression. These results confirm that the proposed model generalizes better across tissue types and yields more reliable segmentation boundaries essential for downstream computational pathology analysis.

**Figure 6.**
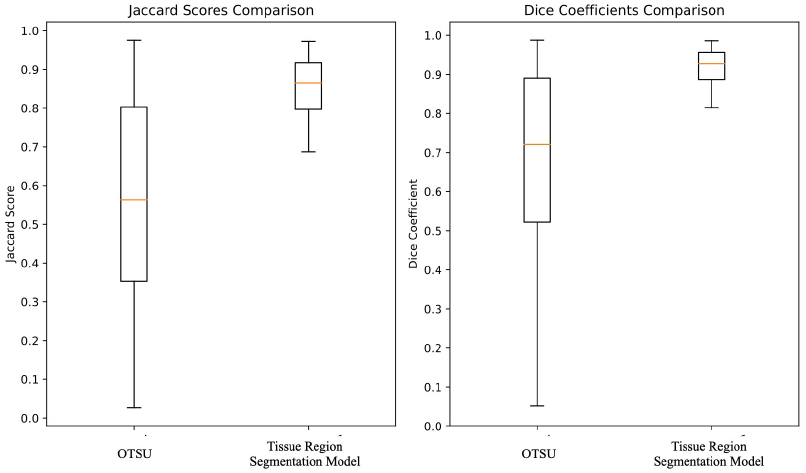
Comparing the tissue region segmentation models’ performances for their generated binary masks and OTSU method using The Jaccard index and Dice coefficients. The tissue segmentation model demonstrates markedly higher consistency and accuracy, with significantly elevated median Jaccard and Dice scores and reduced variability compared to the classical OTSU thresholding method.

Qualitative results (Fig. [7]) show that the proposed model is able to segment the tissue region in faint IHC stained WSIs and H&E stained images with fatty tissue regions, while the mask generated by OTSU, not only segments the tissue as background in almost all images with such characteristics, it segments the artefact with high pixel contrast in WSIs as a tissue region.

**Figure 7.**
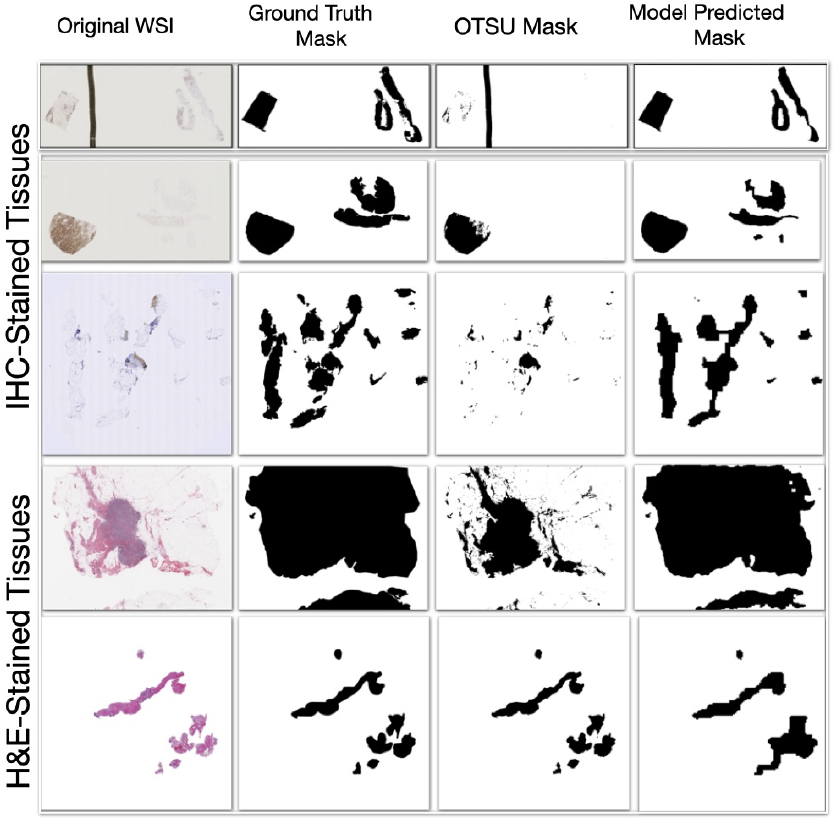
Qualitative comparison of tissue segmentation performance across different stains and tissue types. Columns show the original WSI thumbnails, ground-truth tissue masks, OTSU thresholding baseline, and the proposed CNN model (L4/128 *×* 128). Rows include representative IHC-and H&E-stained slides from various organs (e.g., breast, prostate). The proposed model accurately preserves faint IHC tissue regions while suppressing background over-segmentation frequently observed with OTSU. Notably, in the first row, OTSU incorrectly classifies the dark scanning artefact as tissue due to its color contrast, whereas the proposed model correctly excludes it, demonstrating superior robustness to non-biological structures.

Probability heatmaps (Fig. [8]) confirm that tissue regions are predicted with high confidence (typically ≥ 0.8) and convert cleanly to binary masks and XML overlays for downstream tools.

**Figure 8.**
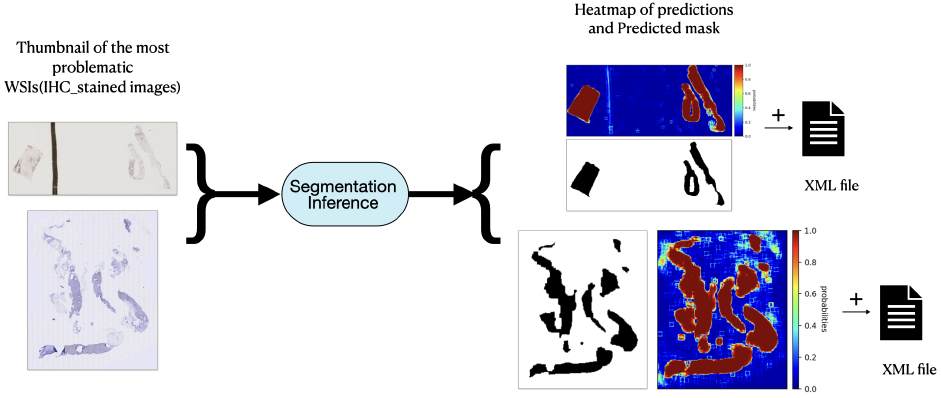
Pipeline view showing probability heatmaps and the derived binary masks. Warmer colors denote higher tissue probability (approaching 1.0). On problematic IHC slides, predictions are high-confidence: heatmaps concentrate near 0.8–1.0 within annotated tissue, and the resulting masks can be exported as XML for downstream tools.

## 5 Discussion

This work demonstrates that accurate and robust WSI tissue-region segmentation can be achieved with a compact CNN operating at *CPU-only* settings. Across diverse cohorts, the proposed model at Level 4 with 128 *×* 128 patches (**L4_128**) consistently performed better comparing to alternative context settings, as evidenced by higher mean F1/AUC and significantly improved paired error profiles. In external validation, the final model which was trained on a combined set of data on different cohorts, achieved **AUC = 0.9988** with 99.01% accuracy, illustrating strong cross-center generalization when training data aggregate multiple stains and institutions. Metrics needed thresholding, reflect a fixed *p* ≥ 0.5 rule across sites; while simple site-specific thresholding/calibration typically restores F1 under domain shift, we did not apply it to preserve a uniform, uncalibrated comparison.

We compared our segmentation results, expressed in terms of the Jaccard index and Dice coefficient, with the findings reported by Bándi *et al*.. Their study utilized FCNN and UCNN to benchmark against the FESI method Bándi et al. [2017] using the same performance metrics. According to their reported results, while the two methods (FCNN and UCNN) do not differ significantly they outperform their traditional counterpart(FESI) with Jaccard index of 0.937 and 0.929 vs. 0.870, p < 0.01. It should be noted that their work did not include a comparison with the OTSU thresholding method.

Our comparison is therefore interpretative and based on published results rather than direct re-implementation. Nonetheless, these cross-study observations emphasize the influence of dataset origin and quality on segmentation outcomes. We observed that a compact CNN can achieve competitive and reliable tissue-region segmentation performance when trained on diverse, high-quality data, even without the architectural complexity of U-Net models such as those used in Bándi et al. [2017]. Our experiments also indicated that while models trained at lower resolutions (Levels 5 and 6) had a high performance, higher-resolution patches consistently yielded superior F1 scores. Furthermore, although large datasets are generally beneficial for deep learning, our results underscore the importance of data quality, variety, and precise annotation in achieving robust generalization across tissue types.

## 6 Conclusion

We demonstrated that a compact, CPU-ready CNN can reliably replace heuristic tissue masking in WSIs, delivering near-ceiling AUC and materially better behavior on challenging stains while remaining simple to deploy. The success of tissue segmentation models in digital pathology imaging is contingent upon a multitude of factors. Paramount among these are the quality and origin of the data, as evidenced by this study’s findings where exhibited superior performance across various tissue types and showcased its ability to handle tissue region segmentation, especially in distinguishing fatty tissues and IHC-stained ones, and their potential utility in practical clinical applications. Moreover, the nuanced characteristics of the samples, such as tissue types, composition, and the inherent complexities of the training process, significantly influence model performance. As digital pathology imaging continues to evolve, refining tissue segmentation models remains an ongoing pursuit. Future advancements hinge on a holistic approach that leverages technological innovation and integrates a comprehensive understanding of tissue intricacies, thus fostering more accurate, reliable, and adaptable models for clinical use.

Limiting the model to only segment the tissue region and background simplify integration but do not address artefact subtypes or multi-class tissue compartments. In future we are planning to extend from binary tissue/background to multi-class masks (artefact strata, tissue borders). Also in future works we are planning to use knowledge distillation which is a representative model compression and acceleration technique where a smaller student model learns from a larger teacher model to help evaluating the efficiency and accuracy trade-offs. Future work should compare the proposed model with distillation-based approaches and broaden benchmarking to include both other traditional image processing techniques rather than OTSU and large-scale deep learning models. Although evaluations were multi-institutional, but broaden cohort diversity (scanners, stains, organs) and release additional masks/splits, where licensing permits, will strengthen generalization and reproducibility in future works.

## Data Availability

The dataset is publicly accessible at https://zenodo.org/records/14131968 Naghshineh Kani et al. [2024c], and the accompanying code is available on https://github.com/sepidehnaghshineh/Tissue-region-segmentation-in-HE-and-IHC-stained-pathology-slides Naghshineh Kani et al. [2024b].

## Acknowledgment

This work was supported by BAU BAP (Grant No. 2022-02.13). Ethical approval for this study was obtained from the Bahçeşehir University Clinical Research Institutional Review Board (IRB approval number: 2022-10/03).

